# Impact of Mass Drug Administration of Ivermectin, Diethylcarbamazine, Albendazole, and Azithromycin on Lymphatic Filariasis, Scabies, and Yaws in West New Britain Province, Papua New Guinea

**DOI:** 10.64898/2026.07.30.26359257

**Authors:** Simon Westby, Joycelyn Salo, Joseph Nale, Wendy Houinei, Jastina Kakul, Michael Payne, Catherine Bjerum, Rose N. Mauyet, Nozomu Aoki, Masato Yamauchi, Susanna Vaz Nery, Magaret C. Baker, Julie Jacobson, Emanuele Giorgi, Moses Laman, Christopher L. King

## Abstract

West New Britain province (WNBP), Papua New Guinea, is the first to combine ivermectin, diethylcarbamazine, albendazole (IDA), and azithromycin for mass drug administration (MDA). We describe the impact of four-drug MDA on lymphatic filariasis (LF), scabies, and yaws through active and passive surveillance.

At baseline, 43 villages were surveyed following selection using model-based geostatistics. Repeat surveys were conducted in 10 high burden villages, one year after MDA. Circulating filarial antigen (CFA) and microfilaria (Mf) microscopy detected LF infections; skin examinations identified yaws and scabies. Electronic data from 27 health centers (HCs) compared monthly attendances in the six months before/after MDA.

Pre-MDA, 151/3,552 (4.3%) participants tested CFA-positive, with 49 (1.4%) Mf-positive. Scabies was identified in 489/3,552 (13.8%) and yaws-like lesions in 130/3,552 (3.6%). In the 10 villages surveyed pre-and post-MDA, CFA positivity decreased by 49% (RR 0.51; CI 0.38, 0.70; p<0.0001; 11.5% to 5.9%), Mf prevalence 85% (RR 0.15; CI 0.07, 0.30; p<0.001; 4.7% to 0.9%), scabies 78% (RR 0.22; CI 0.14 -0.33; p<0.0001; 11.1% to 2.4%) and yaws 31% (RR 0.69; CI 0.41 - 1.14; p=0.15; 3.8% to 2.5%). Mean (SEM) monthly HC attendances for yaws-like lesions decreased from 27.4±0.9 to 15.5±1.6 (p<0.0001) and for other skin diseases from 95.6±3.4 to 72.9±7.7 (p<0.0001), with no significant change in total non-skin disease attendances (728.8±30.9 to 796.8±44.4, p=0.178).

MDA with IDA and azithromycin reduced LF and other skin infections burden in WNBP, demonstrating the effectiveness of four-drug MDA and the potential for using active and passive surveillance to monitor impact.

## INTRODUCTION

Mass drug administration (MDA) is a core public health strategy recommended by the World Health Organization (WHO) to control and eliminate neglected tropical diseases (NTDs), delivering safe and efficacious drugs to entire at-risk populations regardless of individual infection status (1). The rationale is to reduce disease transmission and control morbidity. This approach has been successful in low-resource settings because of its logistical efficiency and cost-effectiveness, often relying on community volunteers to administer drugs donated through large-scale pharmaceutical company programs.

Papua New Guinea (PNG) recently launched an MDA program to eliminate lymphatic filariasis (LF) as a public health problem in line with WHO guidelines, and to control other NTDs that are highly endemic in PNG, including scabies, yaws, and soil-transmitted helminths (STH) (2–7). The current WHO recommendations for LF MDA in areas outside sub-Saharan Africa that have not previously received MDA are to co-administer a single dose of ivermectin, diethylcarbamazine, and albendazole (IDA) twice annually, with coverage of 65% of the population (3, 8). This drug combination is also highly effective against scabies and STH (9, 10). Based on clinical trials, many conducted in PNG, the WHO recommends MDA with azithromycin to eliminate yaws (11). Since LF and yaws are co-endemic in PNG, the PNG National Department of Health (NDOH) has recommended combining IDA with azithromycin for MDA. Recent studies in PNG have established the safety of combining IDA with azithromycin for MDA· (12). In this context, West New Britain province (WNBP) became the first region in PNG to receive MDA with IDA and azithromycin.

One approach to monitor the impact of MDA is passive surveillance through the use of routinely collected data. PNG has recently adopted an electronic National Health Information System (eNHIS) in which health centers (HCs) report monthly data on various diseases including leprosy, yaws-like lesions and other skin disease (undifferentiated). A recent publication using eNHIS data from WNBP suggests a substantial reduction in both yaws and other skin disease HC attendances following the combined MDA (13). However, the uses of electronic health data are constrained by limited record completeness, difficulty in assessing the impact of MDA on health seeking behavior, and disease misclassification, a particular issue for yaws in PNG where several infections cause similar ulcerative disease and serological testing is not routinely available at HCs. At the time of the MDA, the eNHIS did not allow HCs to report LF or scabies, meaning the impact of MDA on these conditions could not be specifically assessed.

Active surveillance, consisting of before-and-after cross sectional surveys, is frequently used to assess the impact of MDA on LF and other NTDs. It involves specifically trained study staff conducting standardized clinical assessments and diagnostic testing at selected sites, thereby reducing susceptibility to information and measurement bias. Active surveillance in sites randomly selected using a population-proportional sampling method is a common approach (14). However, this approach has significant limitations in PNG because LF and other NTDs are more common in rural areas with lower population densities (3), leading to oversampling in urban areas and under sampling in rural ones. Model-based geostatistics (MBG) is an alternative method which uses a mathematical model of spatial correlation to identify sites most likely to harbor LF or other NTDs (15).

Herein, we report the impact of this four-drug combination on the prevalence of LF, yaws and scabies after one round of MDA using active surveillance data collected before and after MDA. We also validate prior eNHIS passive surveillance findings with additional data quality checks and comparison to non-skin disease attendances. We show that one round of MDA with four drugs lowers the prevalence of LF, scabies, yaws, and the number of HC attendances for skin diseases.

## MATERIALS AND METHODS

### Mass Drug Administration

MDA was conducted in December 2023 using a single co-administered dose of ivermectin (200 µg/kg, donated by Merck) and diethylcarbamazine (6 mg/kg, donated by Eisai), based on a validated dosing pole (16), along with a fixed dose of 400 mg of albendazole (donated by GlaxoSmithKline), following WHO guidelines. One week later, individuals received a single oral dose of azithromycin (donated by EMS).

The azithromycin dosing was as follows: persons aged 15 years or older received 2 grams; those aged 10 to 14 years received 1.5 grams; children aged 5 to 9 years received 1 gram; and children aged 1 to 4 years received 500 mg. Since ivermectin is contraindicated in children under 5 years, they received 5% permethrin cream for scabies. All medications were provided free of charge, with direct observation of therapy. An epidemiologic coverage survey measured the proportion of drugs administered relative to the total population. A formal coverage survey was conducted in May 2024 following WHO survey protocols (17). This involved randomly selecting 30 villages. Three villages refused to participate, 27 were surveyed. These included 367 randomly selected households. All household members, regardless of MDA eligibility, were asked to participate. In total, 2,243 individuals were surveyed (1,112 males and 1,130 females), including 852 children aged 2-14 and 1,332 individuals aged 15 and older.

### Cluster selection and sample size

We used MBG to select baseline survey sites likely to have high LF endemicity (2). Evidence suggests that this MBG requires fewer sampling clusters to estimate the overall mean LF prevalence in an area than random sampling (18). The geospatial model utilized available variables associated with LF infection, including elevation, vegetation coverage, distance from the ocean, access to drinking water, window screens, bed nets, and malaria prevalence, since malaria shares the same mosquito vectors as LF in PNG (2). These covariates were used to develop an exceedance probability (EP) score, ranging from 0 to 1, to predict which villages were likely to have LF prevalence exceeding 2%, as previously described (2). All villages with EPs greater than 0.7 were considered for selection, and the final list was confirmed after discussions with provincial officials, with selections constrained by security and seasonal accessibility considerations. Due to security concerns at the time of sampling, sites in the Gloucester region (northwest coast) were excluded from selection. Overall, n=43 villages were sampled (**Figure 1**).

**Figure 1.**
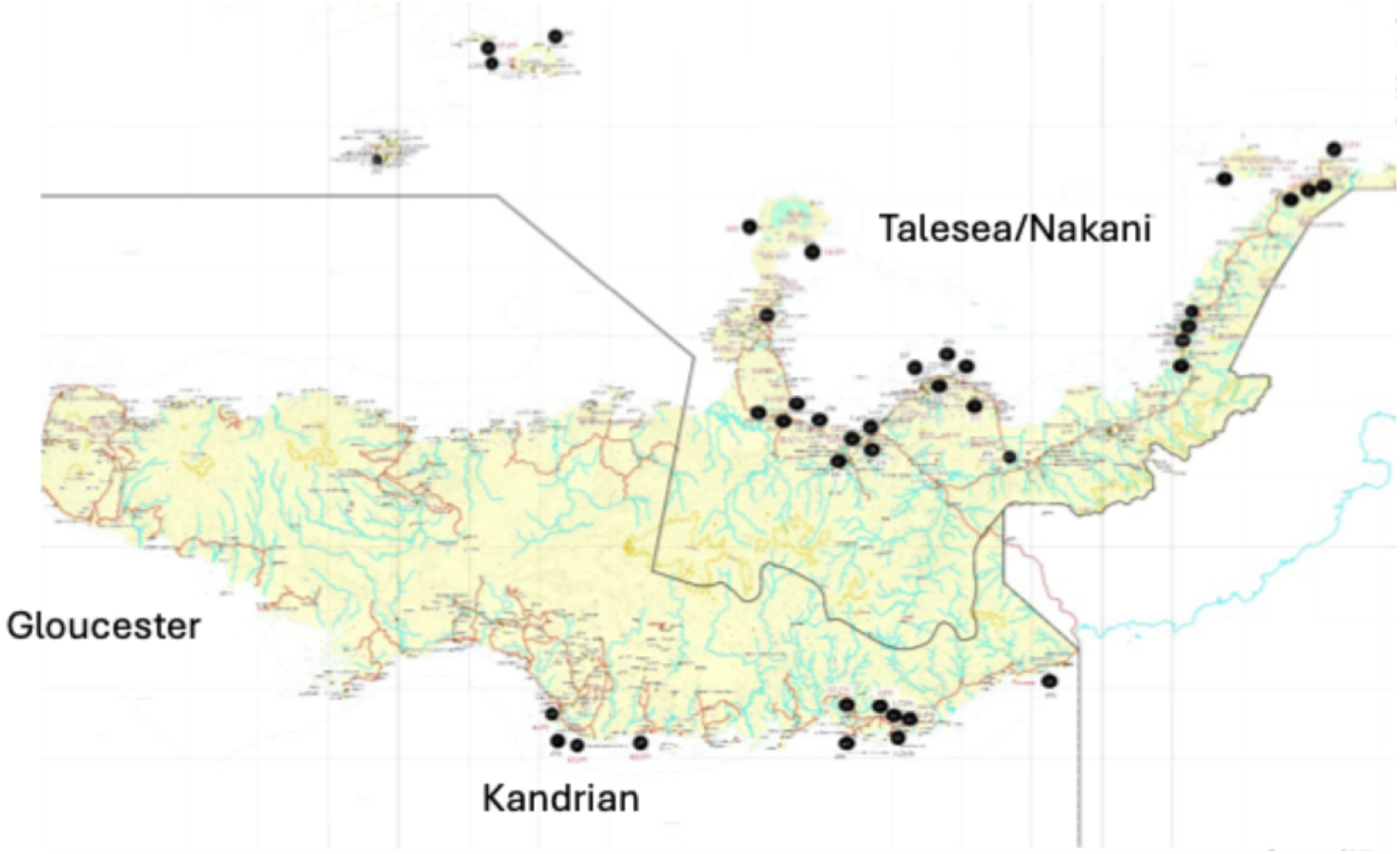
West New Britain Province and location of baseline sampling villages (N=43)

To determine whether there is active LF transmission, WHO sampling guidelines recommend sampling 2,000-3,000 adults across approximately 30 clusters to achieve 95% confidence that there is <1% Mf prevalence in the province, indicating transmission interruption for LF (17). Power increases with the number of villages in the sample to detect Mf <1% (19). Because we aimed to sample children with a higher prevalence of skin diseases but a lower prevalence of LF, the baseline sample size was increased to over 3,000 individuals across 40 villages. The sample consisted of roughly 100 people per cluster (village), with an average household size of 5, and about 20 households were randomly selected per cluster. On the day of the survey, households were selected in each village as follows: first, the village was mapped, and a census was conducted to record all households and their members. The households were then divided into four roughly equal segments, each containing more than 20 households. One segment was randomly chosen from each village, and all individuals over 1 year of age in those households were included in the sample. Baseline sampling ran from September to December 2022. The MDA with IDA and azithromycin took place in December 2023. Post MDA sampling of selected villages occurred from September to November 2024. For village selection post-MDA, all villages that had Mf-positivity at baseline were surveyed. Additional selections were made of villages with the highest baseline prevalences of two or more of: i) circulating filarial antigen (CFA) ii) clinical yaws iii) scabies. A list of fourteen villages was reviewed by the provincial health authorities to account for logistic, weather, and safety parameters, resulting in a final list of ten villages which were sampled.

### Assessment of LF, scabies, and yaws infections

To assess the prevalence of LF, finger-prick blood samples were collected for CFA testing using Filariasis Test Strips (FTS) (Alere, Scarborough, ME, USA) following the manufacturer’s instructions. Tests that did not display a control line were considered invalid and were repeated. The FTS detects a circulating antigen released by *W. bancrofti* (Wb) adult worms into the peripheral blood (20) and acts as a sensitive screening tool for identifying individuals with microfilaremia. Those with positive FTS results then underwent Mf testing using 60-μl thick blood smears prepared from an additional finger-prick sample collected between 9 PM and 1 AM, as previously described (3). Slides were prepared in the field on the day of collection. Where this was not possible, fingerstick blood was collected in containers with anticoagulant, stored in a cooler, agitated every four hours, and transported back to a lab in Kimbe for processing. Two microscopists examined the slides, and in cases of discordant Mf counts, a third reader evaluated the slide. Male participants were asked if they experienced testicular swelling (hydrocele), and all were examined for lymphedema.

To assess the community prevalence of skin diseases, we conducted in-person surveys and skin examinations focused on scabies, yaws, and leprosy for all participants. Personnel responsible for diagnosing skin diseases completed a targeted diagnostic course before data collection. We used the International Alliance for Control of Scabies (IACS) clinical tool for diagnosing scabies (21). This tool is based on four criteria: i) the presence of typical lesions, such as scabies burrows; ii) the typical distribution of the lesions (e.g., the webs of the fingers, hands, wrists, or ankles); iii) the presence of itch; and iv) contact history with an individual with scabies. If an individual met all four criteria, this was recorded as typical scabies. If an individual met three of four criteria, this was classified as suspected scabies. (See **Supplemental Table 1** for a summary of the criteria used for classifying typical and suspected scabies). Pruritic papules with crusting were noted. When diagnostic uncertainty arose, and internet signal permitted, anonymized photos of selected rashes were taken and transmitted to a remote expert for diagnostic assistance.

Yaws-like lesions included typical papules, papillomas, ulcers (usually not painful), and ulceropapillomas. To confirm yaws, we collected additional finger-stick blood samples for the Dual Path Platform (DPP, ChemBio Diagnostic Systems, NY, USA), which detects antibodies to two antigens: a treponemal-specific antigen indicating prior or current yaws or syphilis, and a second non-treponemal antigen that measures non-specific antibodies, similar to the rapid plasma reagin (RPR) antigen. A moderately or strongly positive result correlates with an RPR titer of 1:8 dilution or higher, suggesting active infection (22). This test cannot distinguish between yaws and syphilis; however, skin ulcers on the extremities, especially in children who are not sexually active, are more likely associated with yaws.

Probable leprosy was recorded if the participant had one or more of the following skin features (23): i) Hypopigmented or reddish skin patches (macules) or nodules, with definite and permanent loss of sensation in these patches, particularly to light touch, pain, or temperature. ii) Thickened or enlarged peripheral nerves, with sensory or muscle weakness in the areas supplied by the affected nerves (e.g., ulnar, median, peroneal, or posterior tibial nerves).

### Passive surveillance data collection

HCs maintain a register listing all patients seen and their presumptive diagnoses. Each month, this information is transferred to the eNHIS, which records common illnesses such as respiratory infections, malaria, diarrheal diseases, and skin diseases. The eNHIS allows reporting of three dermatologic conditions: i) yaws-like lesions (yaws), ii) leprosy, and iii) other skin diseases without further details. No other NTDs are included. Before the study, a skin disease training program was conducted for all officers in charge of HCs in WNBP to improve skin disease recording and facilitate the identification and reporting of other skin conditions. The authors helped design and deliver this training in collaboration with NDOH and WHO. Notably, at the conclusion of the study, training sessions in partnership with PNG NDOH have resulted in the inclusion of scabies and lymphedema in eNHIS.

eNHIS records for all WNBP HCs were reviewed from July 2023 to June 2024. HCs with fewer than three months of data in either July–December 2023 or January–June 2024, as well as those that repeatedly entered the same information in two or more consecutive months, were excluded.

### Data acquisition and management

The study used electronic data capture via cell phones. Survey forms were generated in REDCap (v11.0.3, Vanderbilt University) and uploaded to the phones. The forms were completed at enrollment by trained study personnel, including interviews and skin surveys. Test results, such as FTS, DPP, and Mf measurements, were first recorded in logbooks and then transferred to the phones. Data entered on the phones was transmitted to the REDCap server at CWRU daily. A participant key, separate from the REDCap database, was kept at the PNG Institute for Medical Research, linking study ID numbers with personal details like names and birth dates. This participant key was not shared with investigators or staff from Case Western Reserve University. Each week, the data was uploaded to the server and checked for errors. Data were compiled in Microsoft Excel, and statistical analysis was performed using SPSS v29.0.2.0, with graphing done in GraphPad Prism v10.4.0.

### Ethical Approval

The study received ethical approval for the protocol “Applied Research for the Integrated Control and Elimination of Neglected Tropical Diseases in Papua New Guinea” from the PNG Medical Research Council (MRAC 22.17) and the Case Western Reserve University Institutional Review Board (STUDY20220661). Written informed consent was obtained from adults 18 years and older. For individuals who could not read or write, a literate village member witnessed and co-signed the informed consent form. Written informed parental or guardian consent was required for children’s participation, and assent was required for those aged 12 or older.

## RESULTS

**Table 1** shows the baseline population demographics. The median age of 24.8 in the sample is slightly higher than the national median of 22.6 (24). Men are underrepresented, making up a median of 42.5% compared to 52.4% in the overall population (25).

**Table 1.**
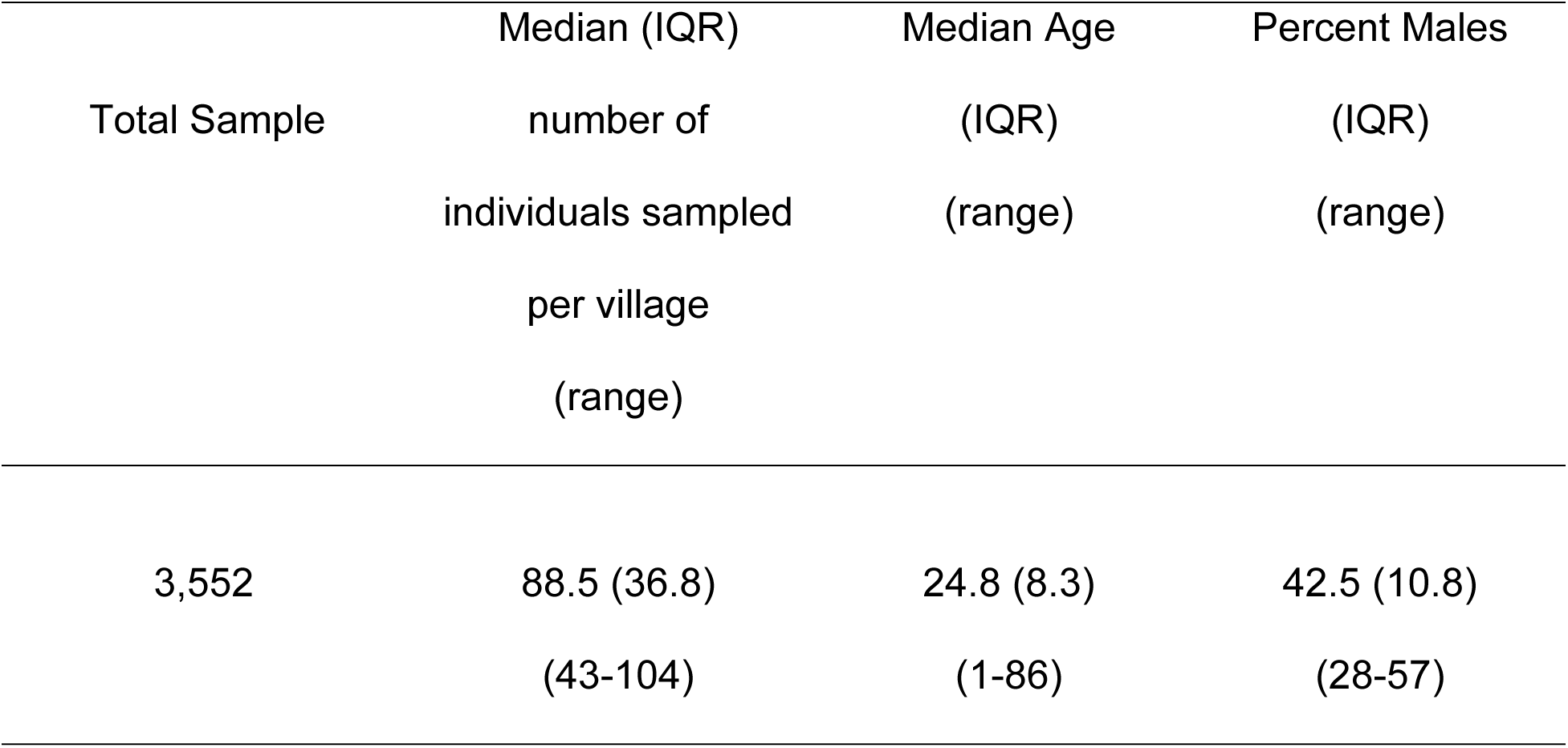
Baseline Population Demographics

Of 3,552 individuals, 151 (4.3; 95% CI 3.6,5.0%) tested positive for CFA. CFA prevalence varied widely across villages, from 0% to 41.4% (**Figures 2 and 3, Supplemental Table 1**). Villages with Mf prevalence >1% or CFA >2% indicate active LF transmission, according to WHO criteria (26). Twelve villages had CFA prevalence exceeding 2%, suggesting ongoing LF transmission. Of the 151 individuals who tested positive on FTS, 147 were tested for Mf, and 49 individuals were Mf positive. Microfilaremia-positive individuals were found in 6 of 43 (14%) villages. The overall Mf prevalence was 1.4% (95% CI 1.0,1.8%; 49 of 3,548), highlighting the highly focal nature of LF infection. One village, Akinam on the south coast, had an Mf prevalence of 33.0% (95% CI 24.3, 42.7%; 36 of 109 subjects). The other five villages had Mf prevalences ranging from 1.8% to 6.0%. Villages with LF-infected participants were clustered in some regions of the province but absent in others (**Figure 3**). Villages within the more populated lower-level government units (LLGs) of Kimbe, Hoskins, and Bialla showed little to no LF, with CFA prevalence below 2% and no Mf-positive individuals (**Figure 3**). On the northeast coast of WNBP in Sule LLG, Nantambu and Noau had CFA prevalences of 3.2% and 15.0%, respectively. Buludava and Kintaku, near Mount Malala at the northern tip of the long peninsula, showed CFA prevalences of 11% and 14.3%, respectively (**Figure 3**). LF prevalence was also high in some villages on WNBP’s south coast in the Kandrian and Gasmata areas. Importantly, sites with CFA prevalence >10% were more likely to have an MBG EP >0.7 (r² = 0.37, p = 0.049), suggesting that MBG may have identified villages more prone to LF.

**Figure 2.**
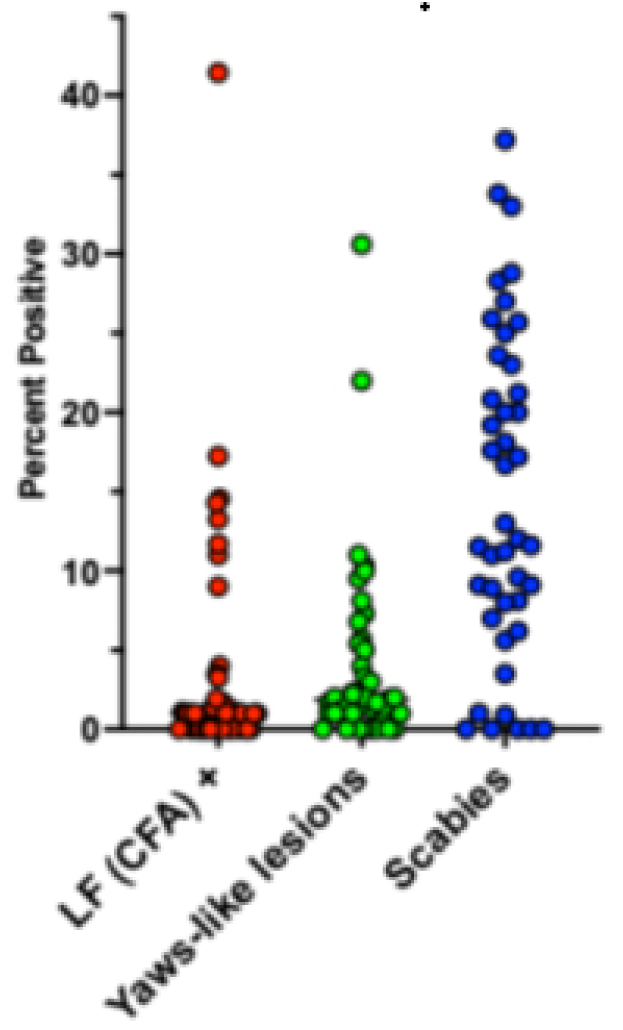
Percent of participants in each of 43 sample villages that were positive for LF (CFA, red), for yaws-like lesions (green), and scabies (black). Each point represents one village.

**Figure 3.**
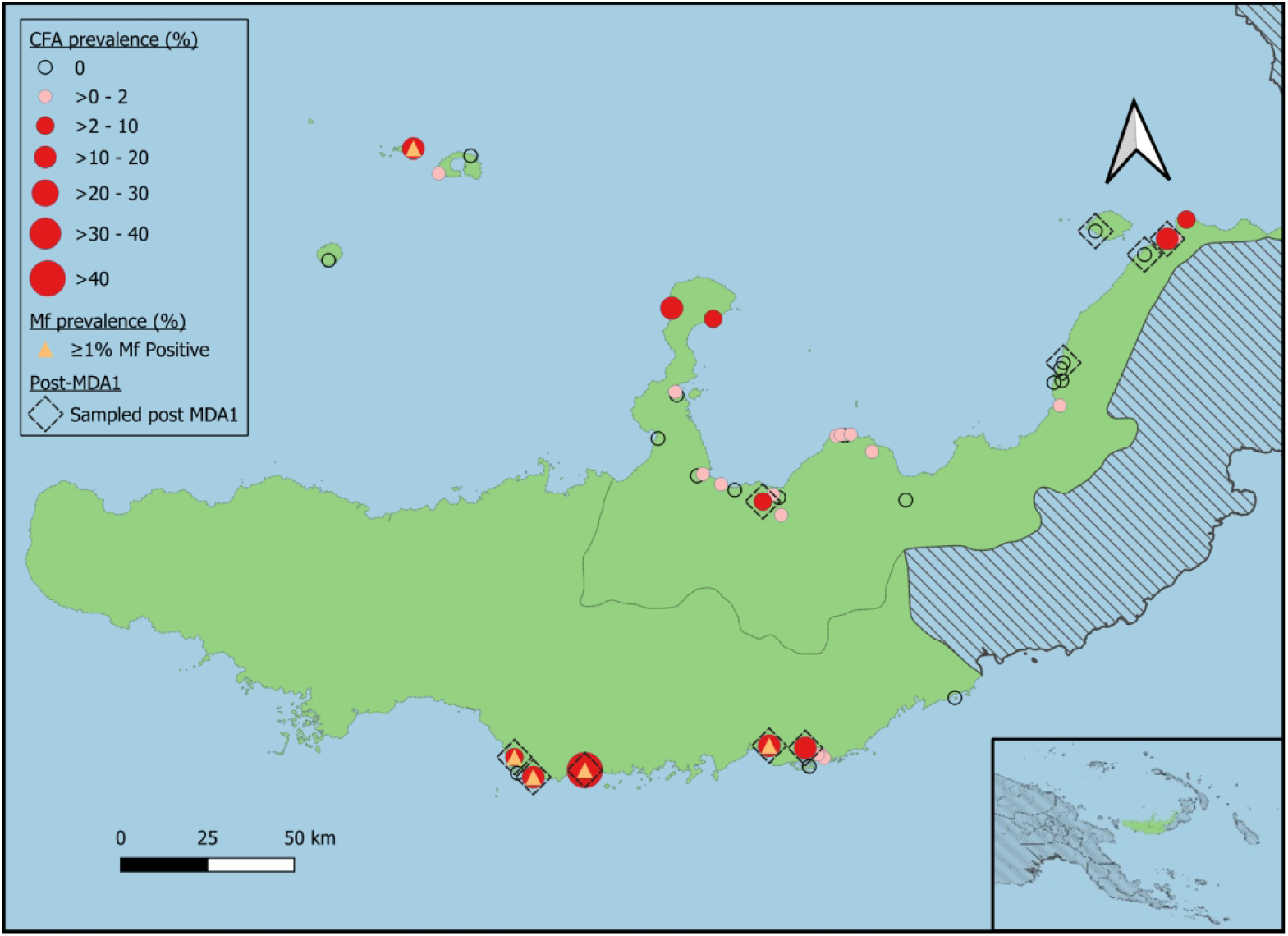
Geographical distribution of LF (CFA) in surveyed villages in WNBP. The size of the red circles indicates LF prevalence in the study villages. Clear circles indicate villages with no FTS-positive individuals, and pink circles indicate villages with FTS positivity of 2% or less. Yellow triangles represent villages with an Mf prevalence of ≥1%. Villages marked with diamonds were resurveyed 1 year after MDA.

CFA levels correlate with the intensity of filarial infection (27). The immunochromatographic FTS test is semiquantitative, and the intensity of the test band correlates with CFA levels (28). FTS scored 1+, 2+, and 3+ with low, moderate, and high CFA levels, respectively, and higher CFA levels are associated with a greater likelihood of being Mf positive (**Table 2**). Ninety percent of individuals with an FTS score of 3+ were also Mf positive.

**Table 2.** Relationship of FTS score and presence and levels of microfilariae

| FTS score | FTS +<br>(N= No. Positive) | Mf + (N)<br>(% Positive) | Mean Mf/ml*<br>(range) |
| --- | --- | --- | --- |
| 1+ | 64 | 8 (13%) | 20 (17-32) |
| **2+** | 31 | 9 (29%) | 38 (17-141) |
| **3+** | 40 | 36 (90%) | 150 (17-1,178) |

\* mean represents those that are Mf positive.

The prevalence and intensity of LF infection increased with age (**Figure 4**). LF prevalence and infection intensity were similar between males and females.

**Figure 4.**
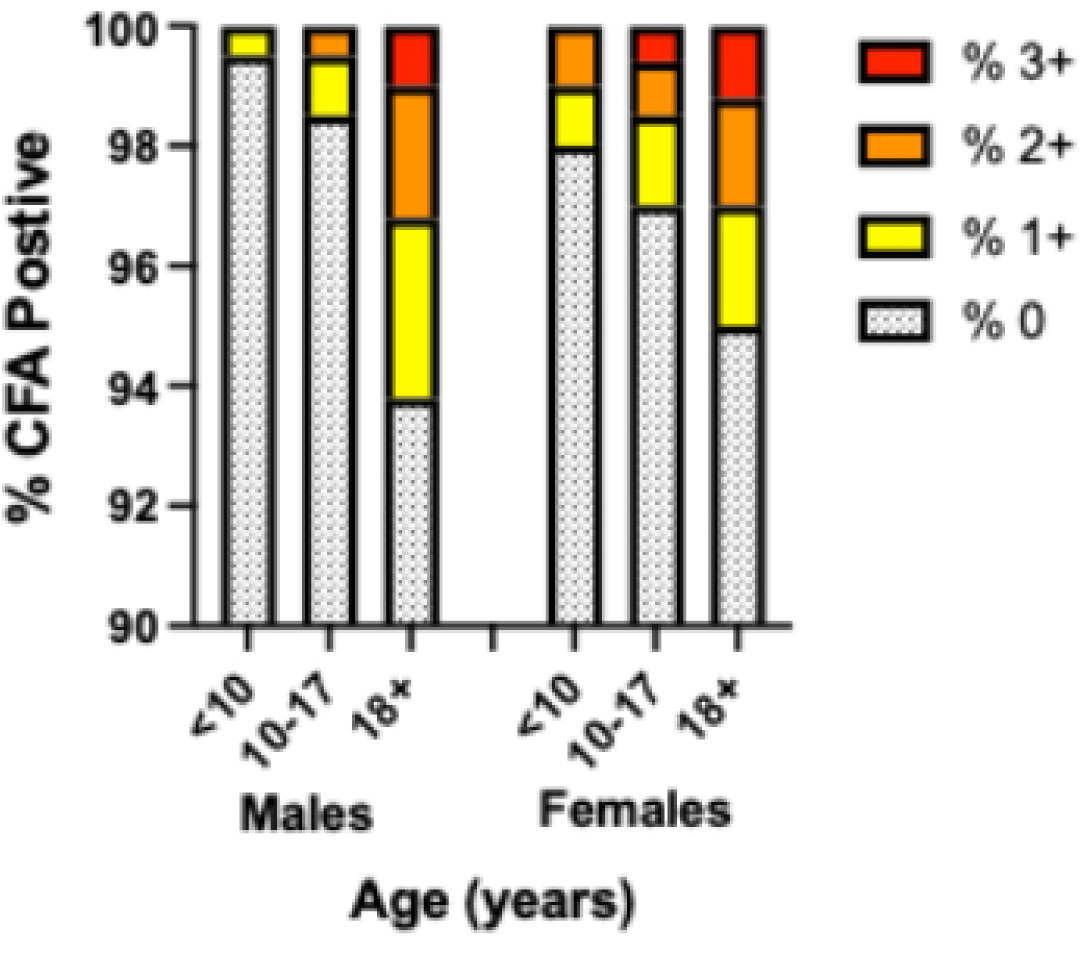
The relationship between age and sex and the prevalence and intensity of LF infection, as determined by the nce and level of circulating filarial antigen (CFA). Unreactive (shaded gray), 1+ (weak, yellow), 2+ (moderate, orange), and by 3+ (strong, red) FTS reactivity

We identified 305 out of 3,552 individuals with typical scabies (8.6%; 95% CI 7.7,9.6%). Additionally, 184 people (5.2%; 95% CI 4.5,6.0%) had scabies-like lesions. Combining both groups, 489 out of 3,540 (13.8%; 95% CI 12.7,14.9%) are likely to have scabies (**Figures 2 and 5, Supplemental Table 3**). The prevalence ranged from undetectable to 37.2% across sentinel sites (**Figures 2 and 5**). Although scabies prevalence varies significantly across study villages, high-prevalence villages are more widespread than those with LF.

**Figure 5.**
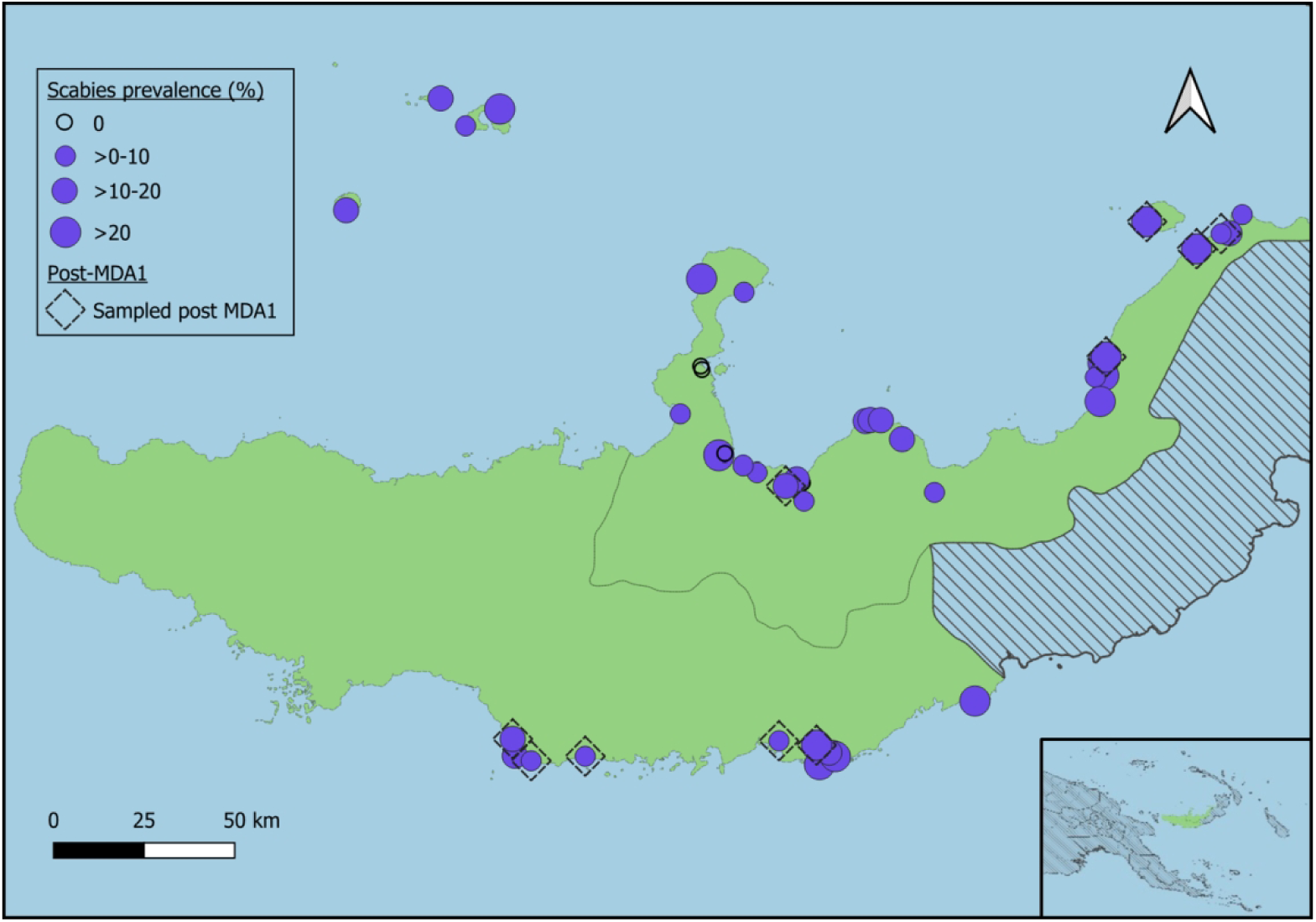
Geographical distribution of scabies (typical and scabies-like) in surveyed villages in WNBP. The size of the purple circles reflects the relative prevalence of scabies in the study villages. Diamonds indicate village that were resurveyed after MDA.

We identified 130 out of 3540 (3.7%, 95% CI 3.1-4.4%) individuals with yaws-like lesions. However, we tested only 40 of the 130 individuals using DPP tests in the field due to limited test availability. Fourteen of the 40 tested (35%) were positive for both bands on the DPP, indicating active yaws. This suggests that 1.3% of the sampled population had active yaws. The prevalence of yaws-like lesions varied among villages (**Figures 2 and 6, Supplemental Table 4),** ranging from undetectable to 30.6%. This uneven distribution of infection prevalence across villages resembled that observed with LF rather than scabies.

**Figure 6.**
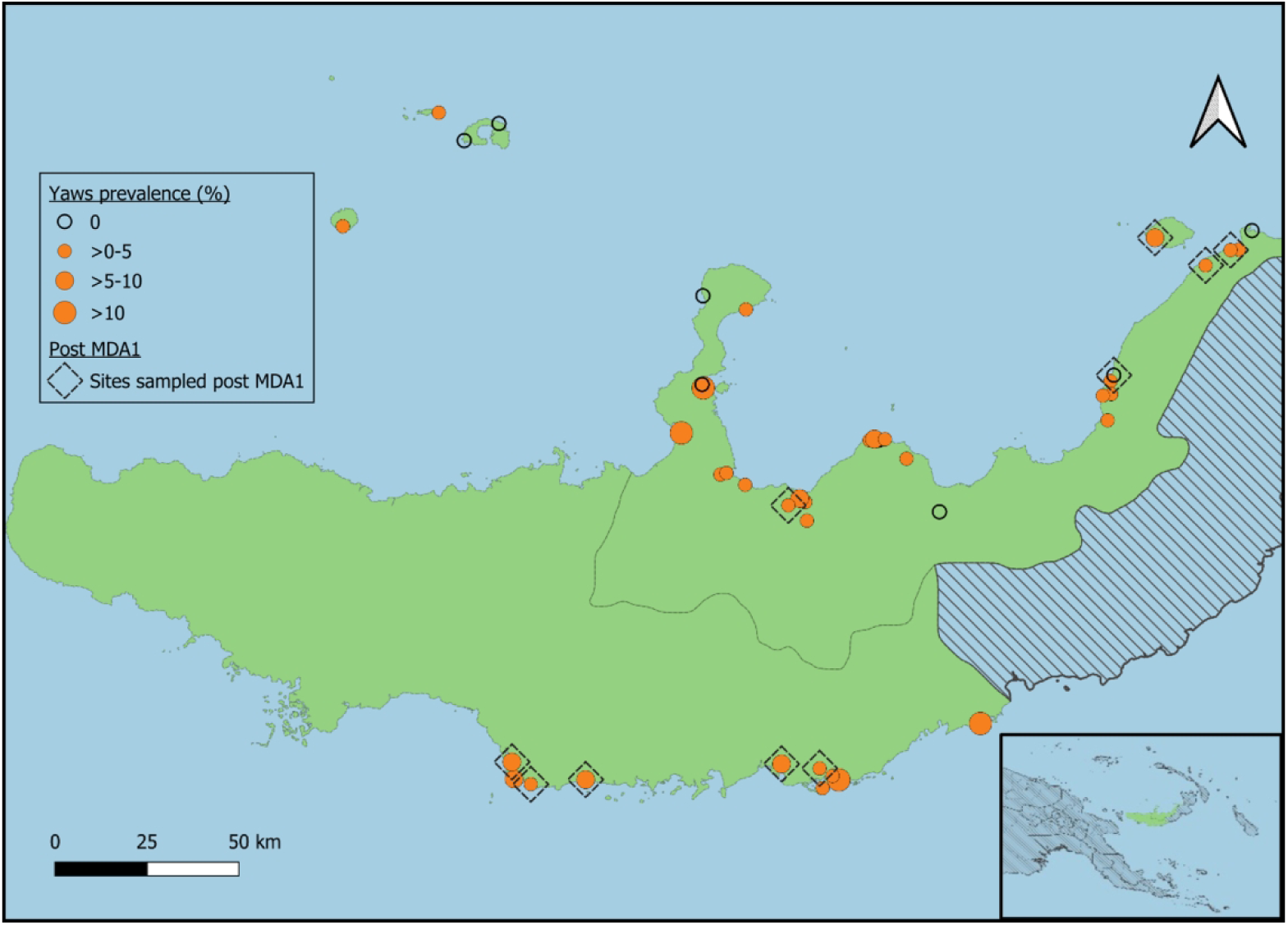
Geographical distribution of yaws-like lesions in surveyed villages in WNBP. The size of the orange circles shows the relative prevalence of scabies in the study villages. Diamonds mark villages that were resurveyed after the MDA.

There was no correlation between the prevalence of yaws, LF and scabies among the 43 villages (r^2^=0.07, 0.01 and 0.125, p>0.05). No cases of leprosy were detected.

### Mass Drug Administration

Epidemiological coverage was reported as 63% for IDA and 55% for azithromycin. The coverage survey found that 74.7% of individuals surveyed were offered the drugs, and 70.1% reported swallowing them. The main reasons for not taking the drugs were absence during distribution or not hearing about the MDA. The primary reason for not swallowing the drugs was fear of side effects. (**Supplemental Table 5**)

### Active surveillance for LF, scabies and yaws-like lesions post-MDA

Ten villages were selected for repeat monitoring and evaluation approximately one year after the MDA (**Figures 3, 5**, and **6** highlighted with diamonds, purple circles in **Figure 8**). In these villages before MDA, 844 individuals were sampled at a median of 88 per village; after MDA, 1038 individuals were sampled, with a median of 110 per village.

After one round of MDA, there was a 49% reduction in CFA prevalence (risk ratio (RR)=0.51; 95% CI 0.38, 0.70; p<0.0001, a prevalence of 11.5% pre-MDA to 5.9% post-MDA) across the 10 sentinel villages, an 85% reduction in Mf prevalence (RR=0.15; 95% CI 0.07,0.30; p<0.0001, from 4.7% to 0.9% prevalence), and a 78% reduction in scabies (RR=0.22; 95% CI 0.14,0.33; p<0.0001, **Figure 7**, from 11.1% to 2.4%). There was a 31% reduction in yaws lesions (RR=0.69; 95% CI 0.41,1.14; p=0.15, from 3.8% to 2.5%), although this change was not statistically significant. Of the 26 individuals with yaws-like lesions observed in the 10 villages post-MDA, 14 were seropositive for treponemal antibodies, but non-treponemal antibody-negative. Five individuals had active yaws (DPP test positive for both treponemal and non-treponemal antibodies), while seven were DPP-negative.

**Figure 7.**
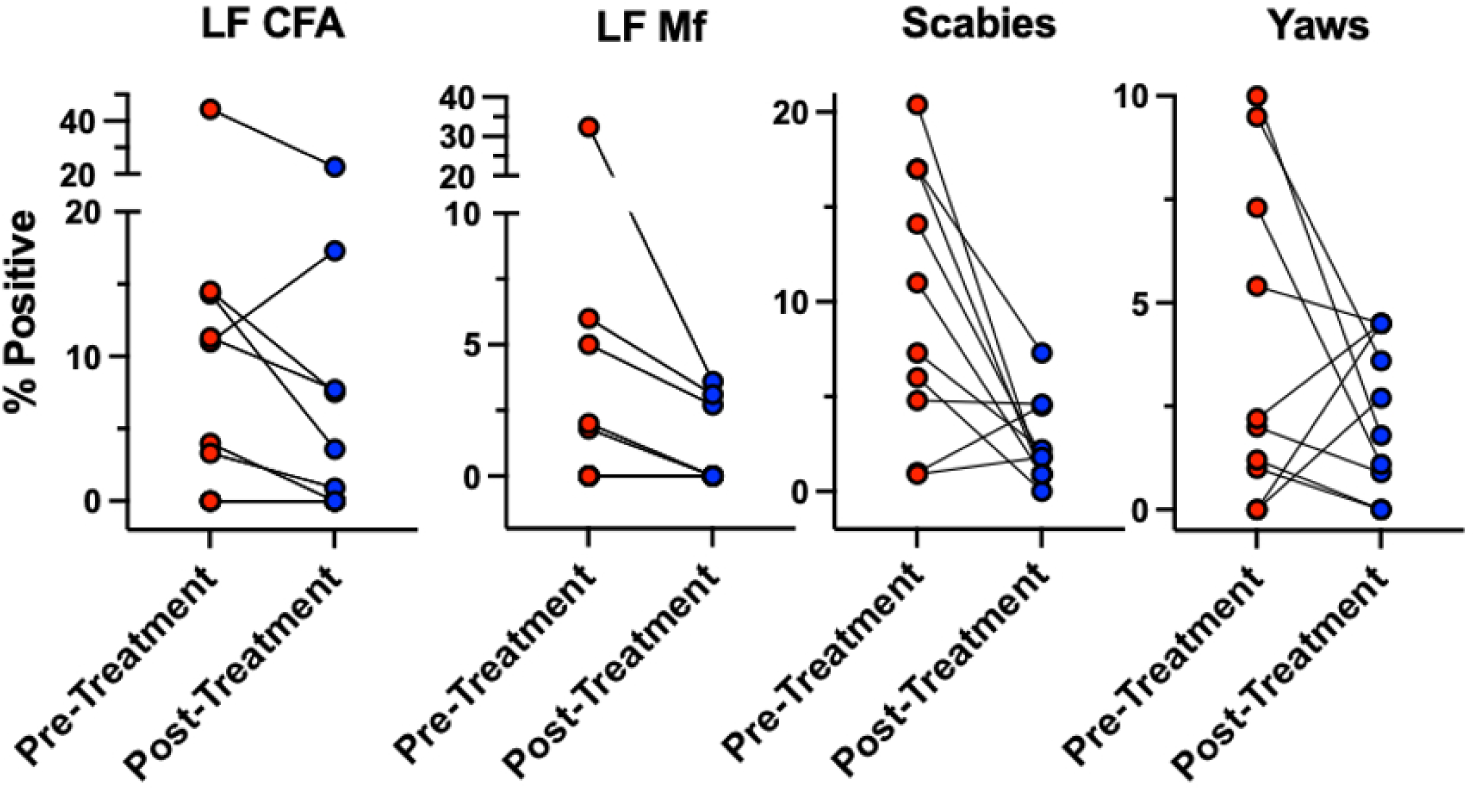
Changes in LF, scabies, and yaws prevalence in 10 villages before and after one round of MDA with IDA followed by azithromycin. The 10 villages are shown in red before MDA and in blue after MDA (N=844 pre - MDA and N=1038 post-MDA).

### Passive surveillance

Data from 27 of the 38 WNBP HCs were included based on pre-defined data quality criteria (**Figure 8**). The mean (standard error of the mean (SEM)) monthly attendance for yaws-like lesions across the 27 clinics prior to MDA was 27.4 ± 0.9 (monthly range 1-122, depending on the month and clinic), which decreased to 15.5 ± 1.6 visits, range 0-103, p<0.0001 (Wilcoxon signed-rank test) (**Figure 9**). Visits for all other non-yaws skin diseases also declined during the same period, from a mean (SEM) of 95.6 ± 3.4 to 72.9 ± 7.7 (p<0.0001). There was no significant change in total visits for non-skin diseases over the same timeframe, with a mean (SEM) of 728.8 ± 30.9 before MDA and 796.8 ± 44.4 after MDA (p=0.178). To account for seasonal variations, we also observed a significant decrease in yaws and other skin disease visits when comparing January-February 2023 pre-MDA to January-February 2024 post-MDA, with yaws-like lesions dropping from 33 ± 3.0 to 15.0 ± 0.8 (p=0.008), and other skin diseases decreasing from 258 ± 30.0 to 60.0 ± 3.7 (p=0.005).

**Figure 8.**
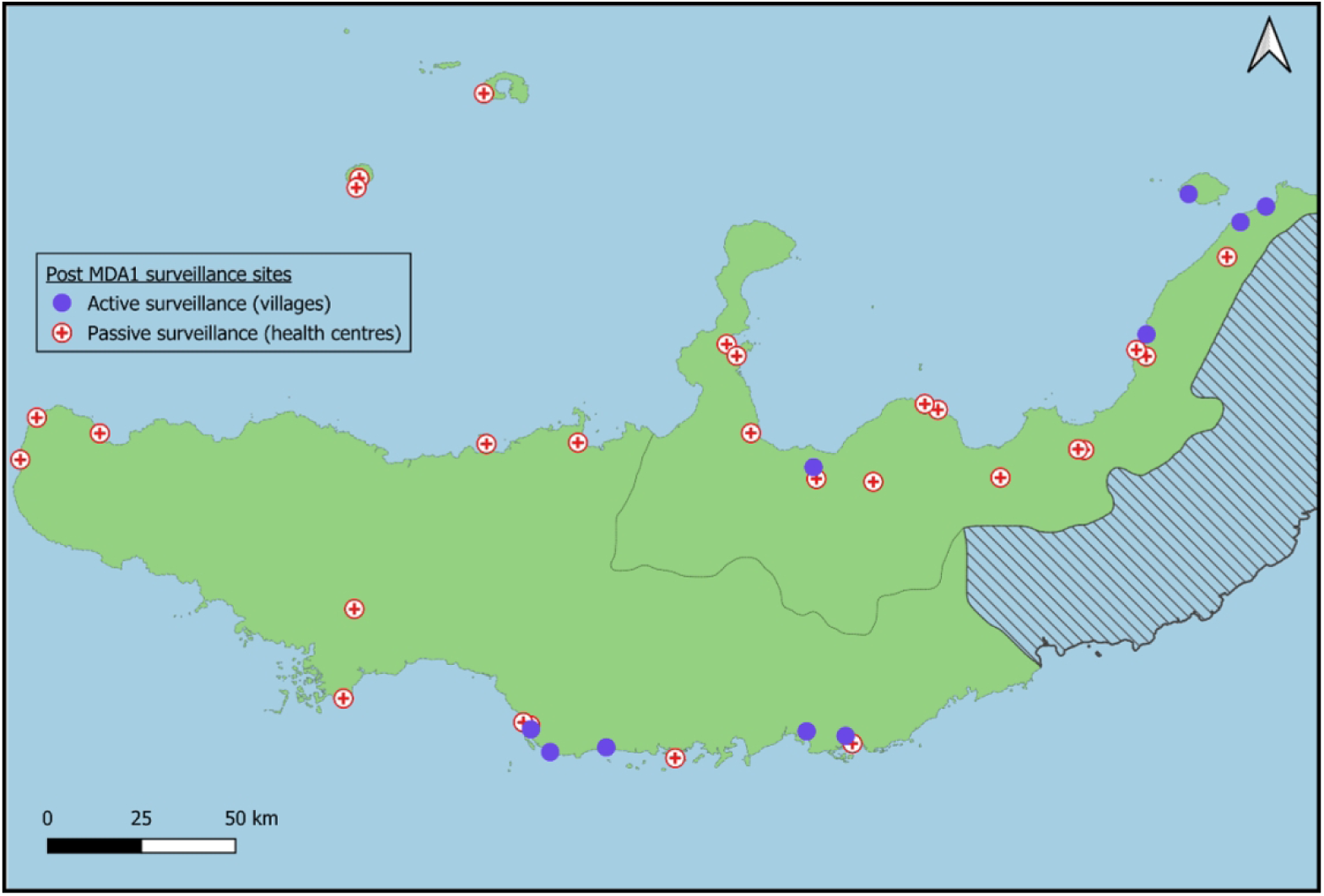
Location of HCs reporting monthly visits for yaw-like lesions and general skin conditions before and after MDA (N=27, red crosses) via the eNHIS. Purple circles represent villages (N=10) that underwent active surveillance for LF, yaws, and scabies before and after MDA.

**Figure 9.**
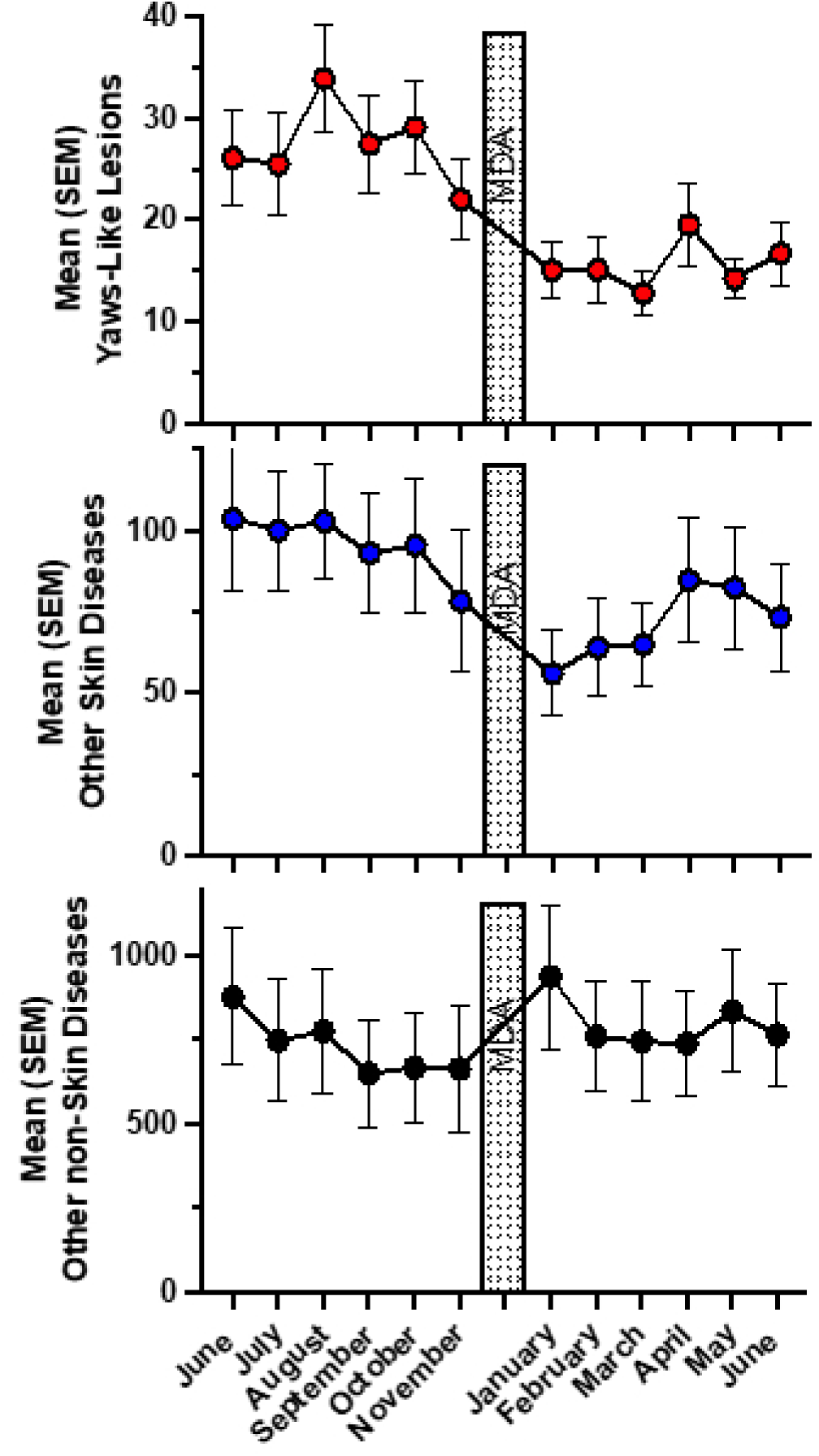
Changes in mean monthly visits at 27 HCs in WNBP, six months before and after MDA for yaw-like lesions (upper panel) other skin diseases (middle panel), and all non-skin diseases (bottom panel). The error bars represent the standard errors of the mean.

## DISCUSSION

PNG is implementing an MDA program including a single co-administered dose of IDA to eliminate LF as a public health problem. Yaws is also highly endemic in PNG, especially in the Island Provinces (4). MDA with azithromycin is the main strategy to eliminate yaws (24, 25). Although these drugs are highly effective against LF and yaws, two or more MDA rounds are recommended to reach elimination. For LF, WHO recommends two annual rounds of IDA with coverage >65% (26); for yaws, three rounds with even higher coverage may be needed, although no specific WHO guidelines have been established (10). PNG is the first country to combine these two MDA programs, administering IDA along with azithromycin. Using data from active and passive surveillance, we observe a substantial decline in LF, scabies, and yaws following a single MDA round.

Active surveillance revealed a 49% reduction in LF CFA prevalence and an 85% reduction in Mf prevalence following MDA, consistent with prior evidence demonstrating IDA’s highly microfilaricidal and more limited macrofilaricidal effectiveness in MDA campaigns (29). We also observed a 78% decrease in scabies prevalence, aligning well with prior studies that have examined the impact of community-wide treatment with ivermectin on scabies prevalence (9, 30).

MDA treatment had a less pronounced impact on yaws, with prevalence reduced by 31%. This likely reflects that azithromycin coverage was lower and some ulcerative conditions reported as yaws may have been due to other infections and thus may have been unresponsive to the treatments administered. In this MDA, azithromycin was given one week after IDA because sufficient safety data for co-administration were not available. This one-week delay likely impacted population adherence to the second drug distribution. Since the study was planned, data—published and unpublished—show that taking all four drugs together has no drug interactions (27) and is safe (11). WHO has approved the co-administration of all four medications simultaneously for future MDA programs in PNG.

We did not observe any cases of leprosy during our active surveillance. This was anticipated given our sample size, that leprosy was eliminated as a public health problem nationally in PNG in 2000, and with contemporaneous data from the PNG National Leprosy Program indicating a leprosy prevalence in WNBP of <1 in 10,000 (31).

Passive surveillance through routine healthcare data has the potential to improve monitoring and evaluation efficiency by reducing site visits, which are costly in PNG due to security, remoteness, and weather, provided that passive surveillance data accurately reflect MDA impact. Passive surveillance using data from the PNG eNHIS shows that MDA significantly reduced visits to HCs for overall skin diseases and yaws-like lesions. Passive surveillance for LF and scabies could not be assessed because these diagnoses were not included in eNHIS at the start of the study. By presenting aggregated monthly reports, we demonstrate the immediate impact of MDA appreciable through the eNHIS; additionally, our data cleaning criteria excluded 28.9% of HCs due to data quality concerns, highlighting the importance of rigorous quality control measures when using PNG routine clinical data. Non-skin disease attendances were not affected by the MDA, suggesting reductions in yaws and other skin disease attendances following MDA were a result of the medications rather than a general phenomenon of reduced health seeking behavior.

One of the study’s goals was to train HC staff in diagnosing various skin diseases, including scabies and lymphedema. These diagnoses are now part of the eNHIS. The eNHIS reported a 24% overall reduction in attendances at HCs for any skin disease in the six months following MDA, aligned with recent findings (13). In rural PNG, the most common skin diseases include scabies, superficial mycoses, pediculosis, eczema, and tropical ulcers (28). Other skin diseases, such as leprosy and lymphedema, occur but at much lower frequencies. Ivermectin treats scabies and pediculosis, while azithromycin treats other bacterial skin infections and tropical ulcers, including yaws. Active surveillance data, which show significant reductions in scabies and LF and a partial reduction in yaws-like lesions, suggest that the overall decrease in skin diseases observed in eNHIS is likely due to the impact of MDA on these infections.

The use of clinical assessments to monitor yaws lesions is challenging because other infections, such as *Haemophilus ducreyi,* can present with similar signs and symptoms and are difficult to distinguish clinically; they also often respond to azithromycin (32). A point-of-care DPP test can detect both specific and non-specific anti-treponemal antibodies, aiding in differentiating yaws from other lesions in pre-sexually active children. Before MDA, only 35% of individuals with yaws-like lesions tested positive by the DPP test, consistent with previous studies on yaws in PNG (11, 33). This suggests that other skin infections or diseases cause most yaws-like lesions detected through physical examination, suggesting passive surveillance alone likely overestimates yaws burden. The overall prevalence of active yaws in WNBP is likely to be between 1.0% and 1.5%, aligning with earlier surveys in PNG (11, 33). Latent yaws may be much higher (34).

This study has several limitations. The baseline survey of WNBP did not include villages from the Gloucester district in the western part of WNBP, although these account for about 20% of the total population. Active surveillance after MDA was conducted in only a few villages, selected based on higher baseline prevalence of the infections studied rather than random sampling. Before MDA, fewer than one in three individuals with yaws-like lesions received a DPP test to confirm yaws. However, more comprehensive DPP testing post-MDA indicates that only about 35% of yaws-like lesions are active cases, although some of these cases may have represented non-yaws lesions in those with latent yaws. Some villages initially selected at baseline for high MBS exceedance probability, and at follow-up due to high baseline burden, could not be sampled because of security and logistical challenges, with similar issues affecting the random selection of households within some villages. Passive surveillance data are inherently limited, as their electronic health records are not collected explicitly for research purposes and are subject to biases, such as misclassification. Data from 27 of the 38 HCs were included in the analysis, as data quality was poor in 11 HCs, potentially introducing selection bias.

Overall, this study is the first to use active and passive surveillance to assess the impact of the IDA-azithromycin combined MDA, emphasizing the safety and effectiveness of this combination and the importance of using both forms of surveillance for monitoring and evaluation, especially in areas like PNG where active surveillance is particularly expensive and time consuming. Integrating these MDA programs in PNG and potentially elsewhere will significantly advance the control and elimination of these widespread and important diseases.

## Data Availability

All data produced in the present study are available upon reasonable request to the authors

## Acknowledgements

We are grateful to the staff at the HCs, the Provincial Health Administration staff, and the many field technicians who made the MDA, monitoring, and evaluation possible. We are grateful to Grace Michael in WNBP for her efforts during field work and for Mary Yohogu for her efforts at the National Department of Health in implementing the MDA. Many thanks to the study participants across WNBP.

## Conflicts of Interest

### Financial Support

Takeda Pharmaceuticals provided major funding for the project through the non-profit organization Bridges for Development. Funding was also provided by a Partnership grant from the Australian NMHRC grant RG204078.

### Author Roles

The project was conceived by JJ, CLK, MCB, KS, SW, JN, ML, and SvN. JJ and SvN secured the funding. LJ, SJ, WH, MY, JN, JK, RN, NA, and MY were responsible for MDA, and NA and MY conducted the coverage survey. EG assisted in selecting villages using model-based geostatistics. JS, SW, ML, MP, CB, and CLK handled baseline and follow-up monitoring and surveillance. CLK and SW analyzed the data and drafted the initial version, while JN, SvN, JJ, and MCB contributed input on later drafts.

### Figure Alt Text

Figure 1. Map of West New Britain province, Papua New Guinea, using black dots to indicate villages which were sampled at baseline as part of monitoring and evaluation prior to the first round of mass drug administration. Shows villages sampled were in Talasea/Nakanai and Kandrian districts with no villages sampled in Gloucester.

Figure 2. Graph showing the range of percentage of people in each village sampled at baseline who were positive for lymphatic filariasis, yaws-like lesions and/or scabies.

Figure 3. Map of West New Britain province, Papua New Guinea showing the range prevalence of lymphatic filariasis circulating filarial antigen and microfilaria positivity in sites sampled prior to the first mass drug administration and indicating which sites were resampled after the mass drug administration. Five of the villages had microfilarial prevalence ≥1%.

Figure 4. Graph of lymphatic filariasis circulating filarial antigen positivity by sex (male and female), each grouped by age (<10 years old, 10-17 and 18+). Circulating filarial antigen positivity is divided into 0, 1+, 2+ and 3+. As age increases, the proportion of 2 and 3+ increases for both genders.

Figure 5. Map of West New Britain province, Papua New Guinea showing the prevalence of scabies in sites sampled prior to the first mass drug administration and indicating which sites were resampled after the mass drug administration. The prevalence of scabies varied across sampled sites.

Figure 6. Map of West New Britain province, Papua New Guinea showing the prevalence of yaws in sites sampled prior to the first mass drug administration and indicating which sites were resampled after the mass drug administration. The prevalence of yaws varied across the sampled sites.

Figure 7. Four graphs showing prevalence of lymphatic filariasis circulating filarial antigen positivity, microfilaria positivity, scabies and yaws pre and post mass drug administration across the ten sites sampled pre and post mass drug administration, showing a reduction across all pairings.

Figure 8. A map of West New Britain province, Papua New Guinea, showing the ten villages actively sampled and the 27 health centres included in passive surveillance following the first round of mass drug administration. The health centres were spread across all three districts.

Figure 9. Three graphs showing monthly mean attendances for yaws-like lesions, other skin diseases and non-skin diseases across the 27 health centres in the 6 months before and 6 months after the first round of mass drug administration, showing a reduction in yaws-like lesions and other skin disease attendances but not in non-skin disease attendances.

### Authors current contact information

Simon Westby - Papua New Guinea Institute of Medical Research, Homate Street, Goroka, Eastern Highlands Province, 441, Papua New Guinea

Joycelyn Salo - Papua New Guinea Institute for Medical Research, Goroka, EHP, Papua New Guinea. j

Joseph Nale - West New Britain Provincial Health Authority, Executive Management, WNBP, Papua New Guinea. j

Wendy Hoiunei - National Department of Health, Port Moresby, Papua New Guinea.

Jastina Kakul - West New Britain Provincial Health Authority, Public Health, WNBP,

Papua New Guinea.

Christopher L. King - Center for Global Health and Diseases, Case Western Reserve

University School of Medicine, OH, USA.

Michael Payne - Center for Global Health and Diseases, Case Western Reserve University School of Medicine, OH, USA.

Catherine Bjerum - Center for Global Health and Diseases, Case Western Reserve University School of Medicine, OH, USA.

Rose N. Mauyet - The Project for Elimination of Lymphatic Filariasis Phase II, Japan International Cooperation Agency, Papua New Guinea.

Nozomu Aoki - The Project for Elimination of Lymphatic Filariasis Phase II, Japan International Cooperation Agency, Papua New Guinea.

Masato Yamauchi - The Project for Elimination of Lymphatic Filariasis Phase II, Japan International Cooperation Agency, Papua New Guinea.

Susanna Vaz Nery - The Kirby Institute, UNSW Sydney, New South Wales, Australia. Magaret Baker - Georgetown University, Washington D.C, USA.

Julie Jacobson - Bridges to Development, Vashon, WA, USA.

Emanuele Giorgi - CHICAS, Lancaster Medical School, Lancaster University, Lancaster, UK.

Moses Laman - Papua New Guinea Institute for Medical Research, Goroka, EHP, Papua New Guinea.

Christopher L. King - Center for Global Health and Diseases, Case Western Reserve University School of Medicine, OH, USA.

## Supplemental materials

**S1.**
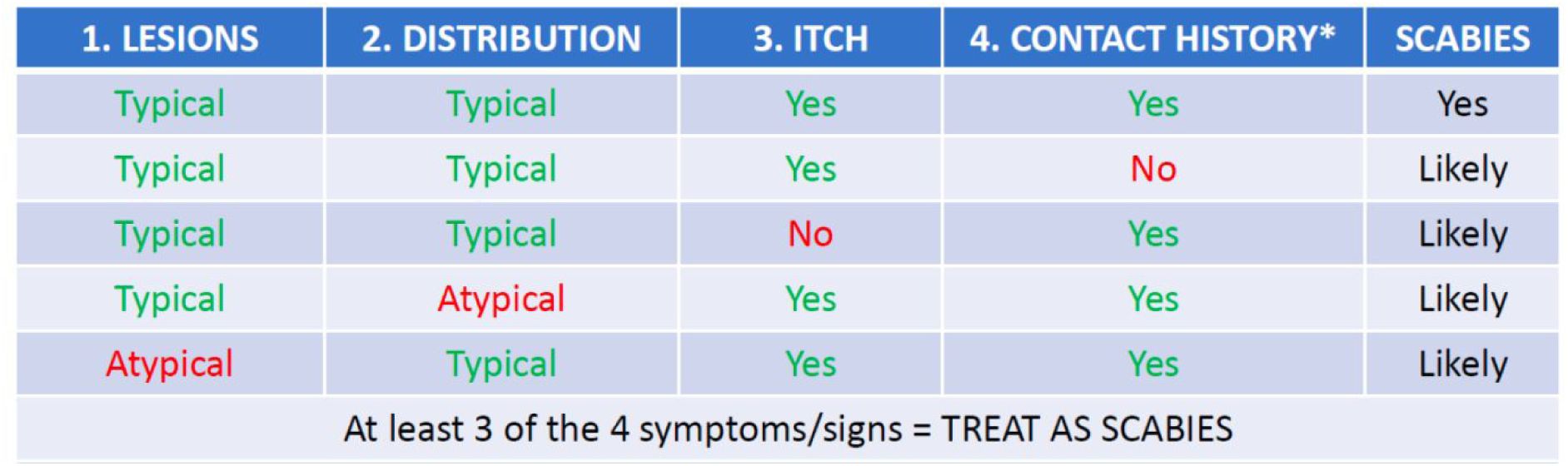
Criteria used by field staff to determine whether individuals had scabies or not.

| 1. LESIONS | 2. DISTRIBUTION | 3. ITCH | 4. CONTACT HISTORY* | SCABIES |
| --- | --- | --- | --- | --- |
| Typical | Typical | Yes | Yes | Yes |
| Typical | Typical | Yes | No | Likely |
| Typical | Typical | No | Yes | Likely |
| Typical | Atypical | Yes | Yes | Likely |
| Atypical | Typical | Yes | Yes | Likely |
| At least 3 of the 4 symptoms/signs = TREAT AS SCABIES |  |  |  |  |

**S2.**
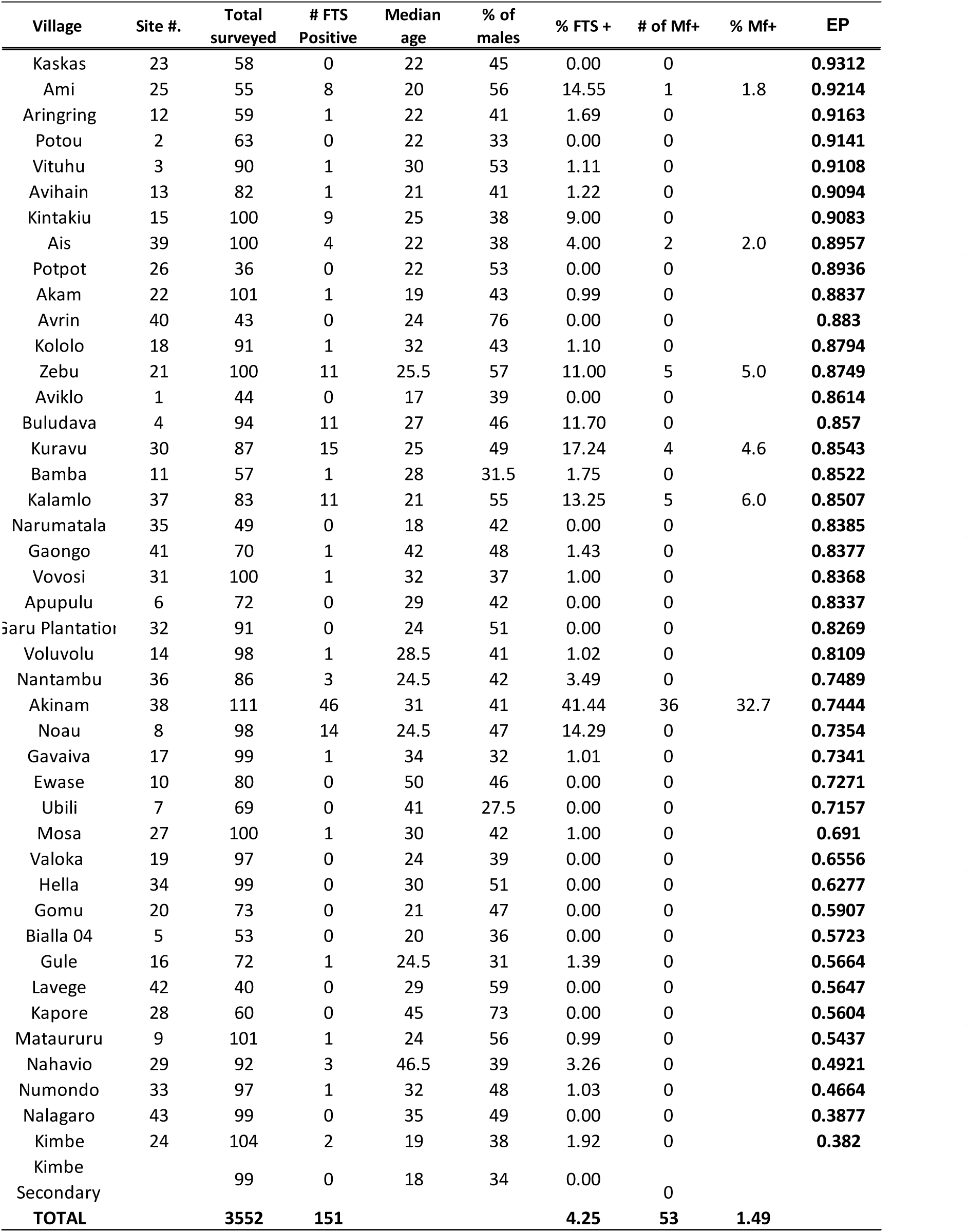
Baseline village demographics, LF parameters and exceedance probabilities (EP)

**S3.**
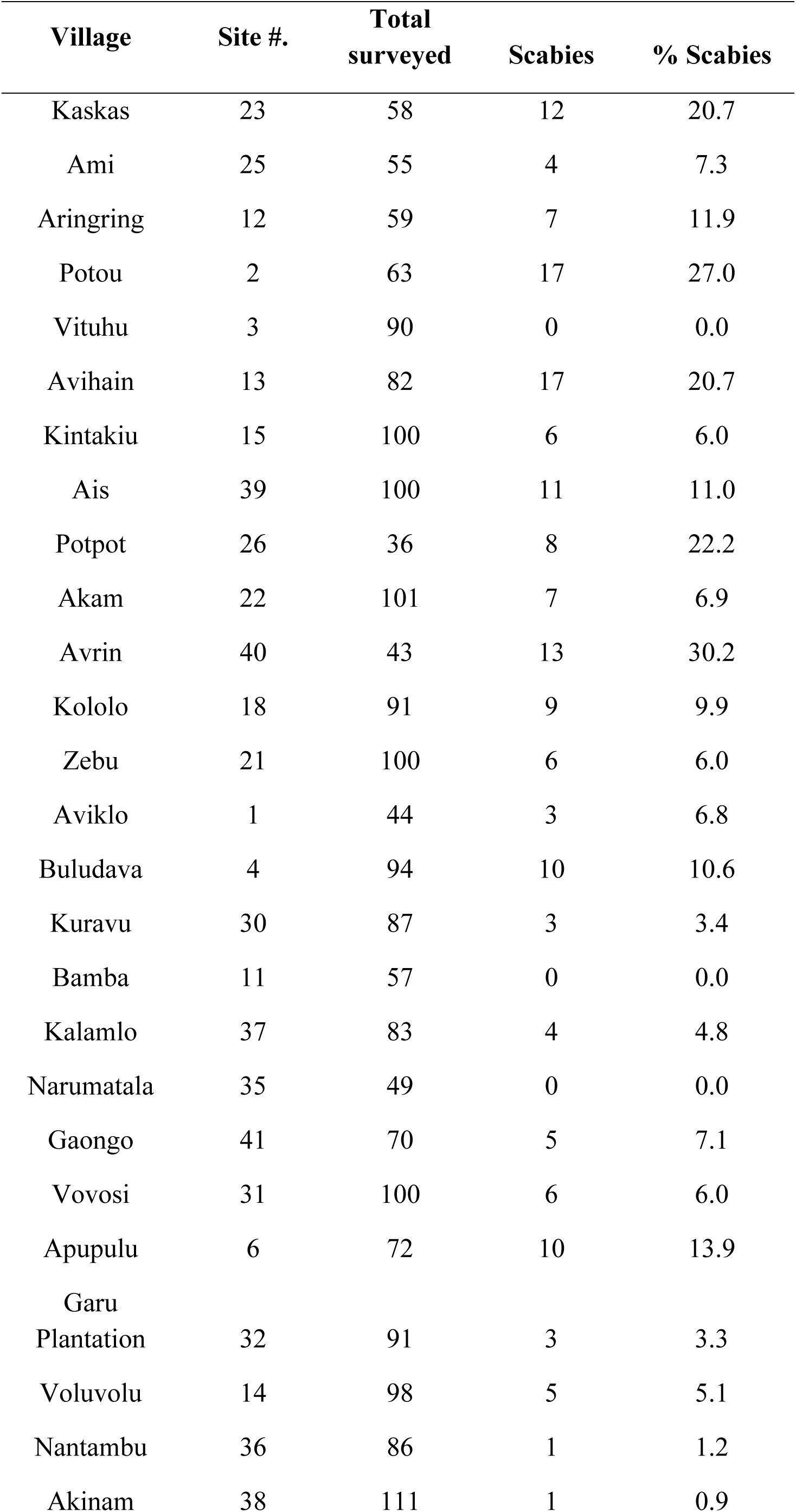
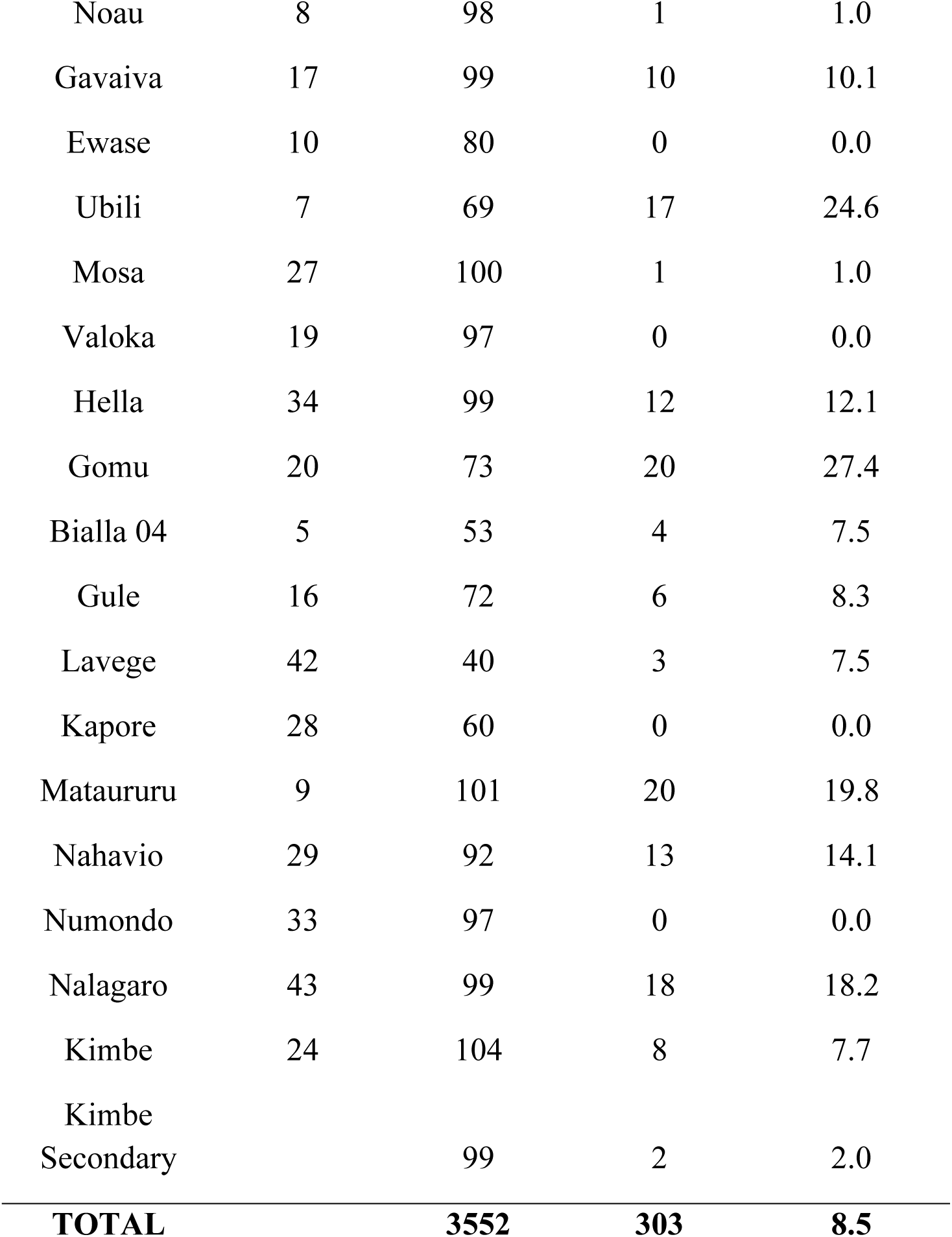
Baseline village scabies prevalence.

**S4.**
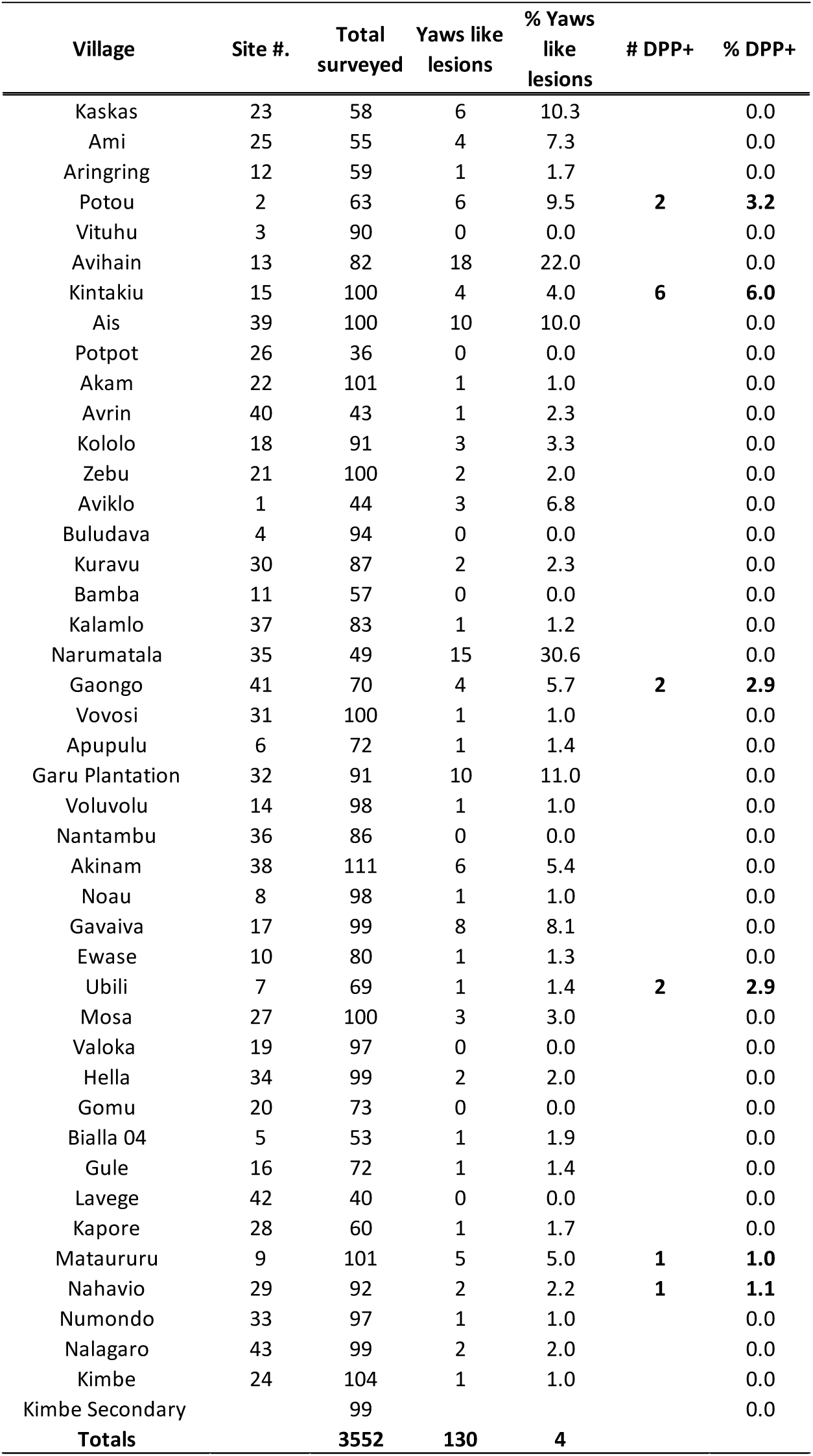
Baseline village yaws prevalence.

**S5.** Official coverage survey results after MDA 1, WNBP.

S5 – Official coverage survey results after MDA 1, WNB
|  | DEC |  | ALB |  | IVM |  | AZM |  |
| --- | --- | --- | --- | --- | --- | --- | --- | --- |
|  | n | % | n | % | n | % | n | % |
| Number of responses | 2,241 |  | 2,236 |  | 2,236 |  | 2,240 |  |
| MDA drugs received | 1,676 | 74.8 | 1,680 | 75.1 | 1,683 | 75.3 | 1,674 | 74.7 |
| MDA drugs consumed among those provided with MDA medicines | 1,575 | 93.8 | 1,585 | 94.3 | 1,591 | 94.2 | 1,566 | 93.3 |
| **MDA coverage** |  | **70.3** |  | **70.9** |  | **71.2** |  | **69.9** |

