## Supplemental Materials for "Impact of Mass Drug Administration of Ivermectin, Diethylcarbamazine, Albendazole, and Azithromycin on Lymphatic Filariasis, Scabies, and Yaws in West New Britain Province, Papua New Guinea"

**Contents**

S1 – Criteria used by field staff to determine whether individuals had scabies or not………..2

S2 – Baseline village demographics, LF parameters and exceedance probabilities (EP)……..3

S3 – Baseline village scabies prevalence .............................………………………………….4

S4 – Baseline village yaws prevalence……………………………………………………………………………………..6

S5 – Official coverage survey results after MDA 1, WNBP…..………………………………8

**S1 – Criteria used by field staff to determine whether individuals had scabies or not**

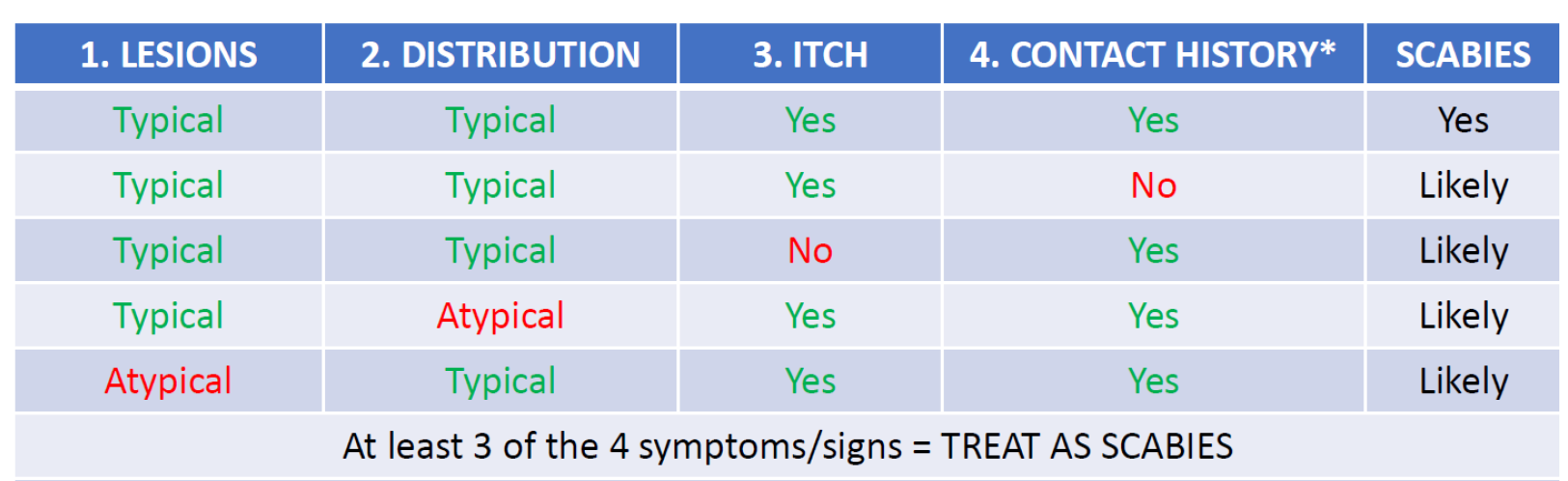

**S2 – Baseline village demographics, LF parameters and exceedance probabilities (EP)**

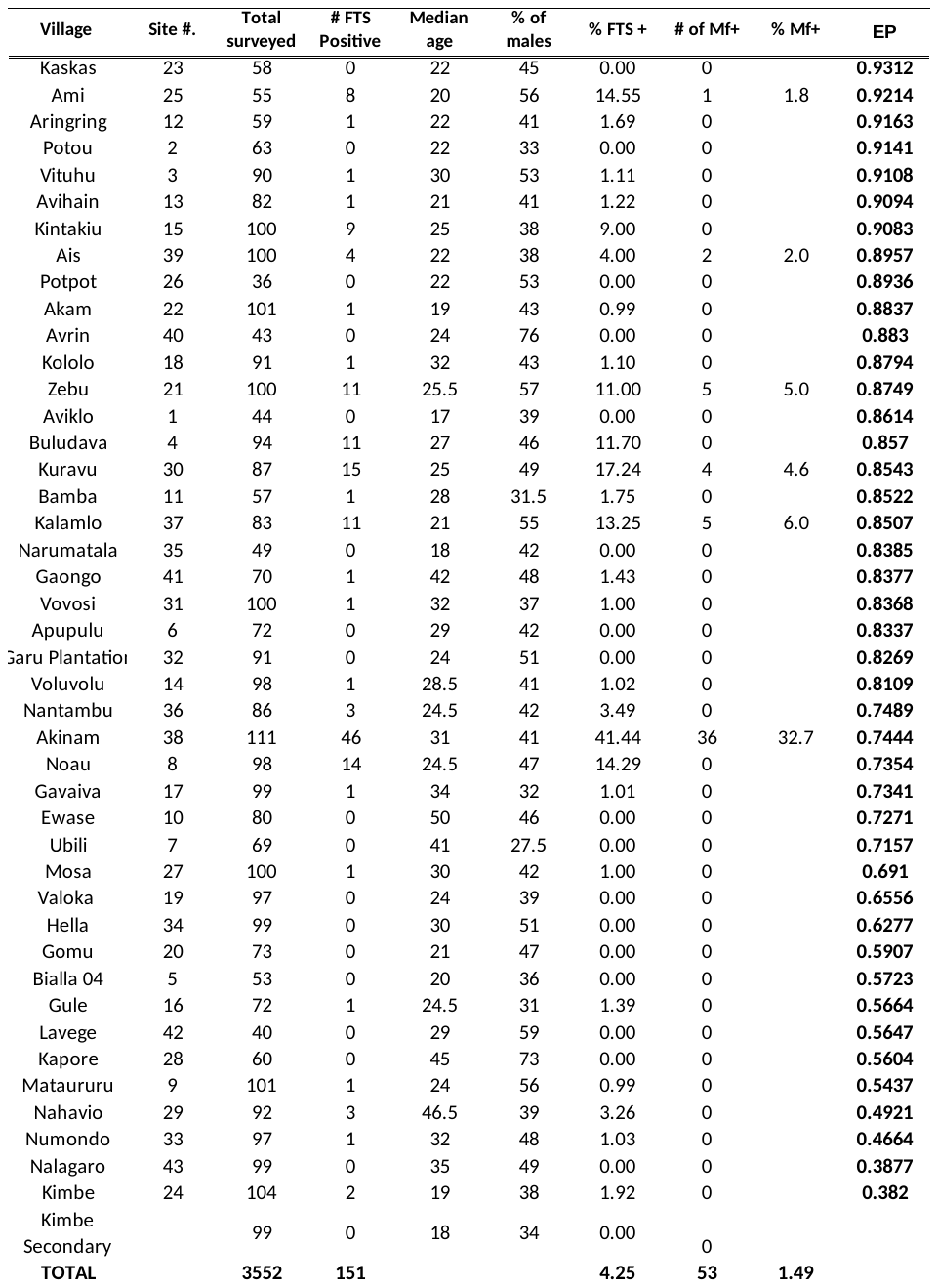

**S3 – Baseline village scabies prevalence**

| **Village** | **Site #.** | **Total surveyed** | **Scabies** | **% Scabies** |
| --- | --- | --- | --- | --- |
| Kaskas | 23 | 58 | 12 | 20.7 |
| Ami | 25 | 55 | 4 | 7.3 |
| Aringring | 12 | 59 | 7 | 11.9 |
| Potou | 2 | 63 | 17 | 27.0 |
| Vituhu | 3 | 90 | 0 | 0.0 |
| Avihain | 13 | 82 | 17 | 20.7 |
| Kintakiu | 15 | 100 | 6 | 6.0 |
| Ais | 39 | 100 | 11 | 11.0 |
| Potpot | 26 | 36 | 8 | 22.2 |
| Akam | 22 | 101 | 7 | 6.9 |
| Avrin | 40 | 43 | 13 | 30.2 |
| Kololo | 18 | 91 | 9 | 9.9 |
| Zebu | 21 | 100 | 6 | 6.0 |
| Aviklo | 1 | 44 | 3 | 6.8 |
| Buludava | 4 | 94 | 10 | 10.6 |
| Kuravu | 30 | 87 | 3 | 3.4 |
| Bamba | 11 | 57 | 0 | 0.0 |
| Kalamlo | 37 | 83 | 4 | 4.8 |
| Narumatala | 35 | 49 | 0 | 0.0 |
| Gaongo | 41 | 70 | 5 | 7.1 |
| Vovosi | 31 | 100 | 6 | 6.0 |
| Apupulu | 6 | 72 | 10 | 13.9 |
| Garu Plantation | 32 | 91 | 3 | 3.3 |
| Voluvolu | 14 | 98 | 5 | 5.1 |
| Nantambu | 36 | 86 | 1 | 1.2 |
| Akinam | 38 | 111 | 1 | 0.9 |
| Noau | 8 | 98 | 1 | 1.0 |
| Gavaiva | 17 | 99 | 10 | 10.1 |
| Ewase | 10 | 80 | 0 | 0.0 |
| Ubili | 7 | 69 | 17 | 24.6 |
| Mosa | 27 | 100 | 1 | 1.0 |
| Valoka | 19 | 97 | 0 | 0.0 |
| Hella | 34 | 99 | 12 | 12.1 |
| Gomu | 20 | 73 | 20 | 27.4 |
| Bialla 04 | 5 | 53 | 4 | 7.5 |
| Gule | 16 | 72 | 6 | 8.3 |
| Lavege | 42 | 40 | 3 | 7.5 |
| Kapore | 28 | 60 | 0 | 0.0 |
| Mataururu | 9 | 101 | 20 | 19.8 |
| Nahavio | 29 | 92 | 13 | 14.1 |
| Numondo | 33 | 97 | 0 | 0.0 |
| Nalagaro | 43 | 99 | 18 | 18.2 |
| Kimbe | 24 | 104 | 8 | 7.7 |
| Kimbe Secondary |  | 99 | 2 | 2.0 |
| **TOTAL** |  | **3552** | **303** | **8.5** |

**S4 – Baseline village yaws prevalence**

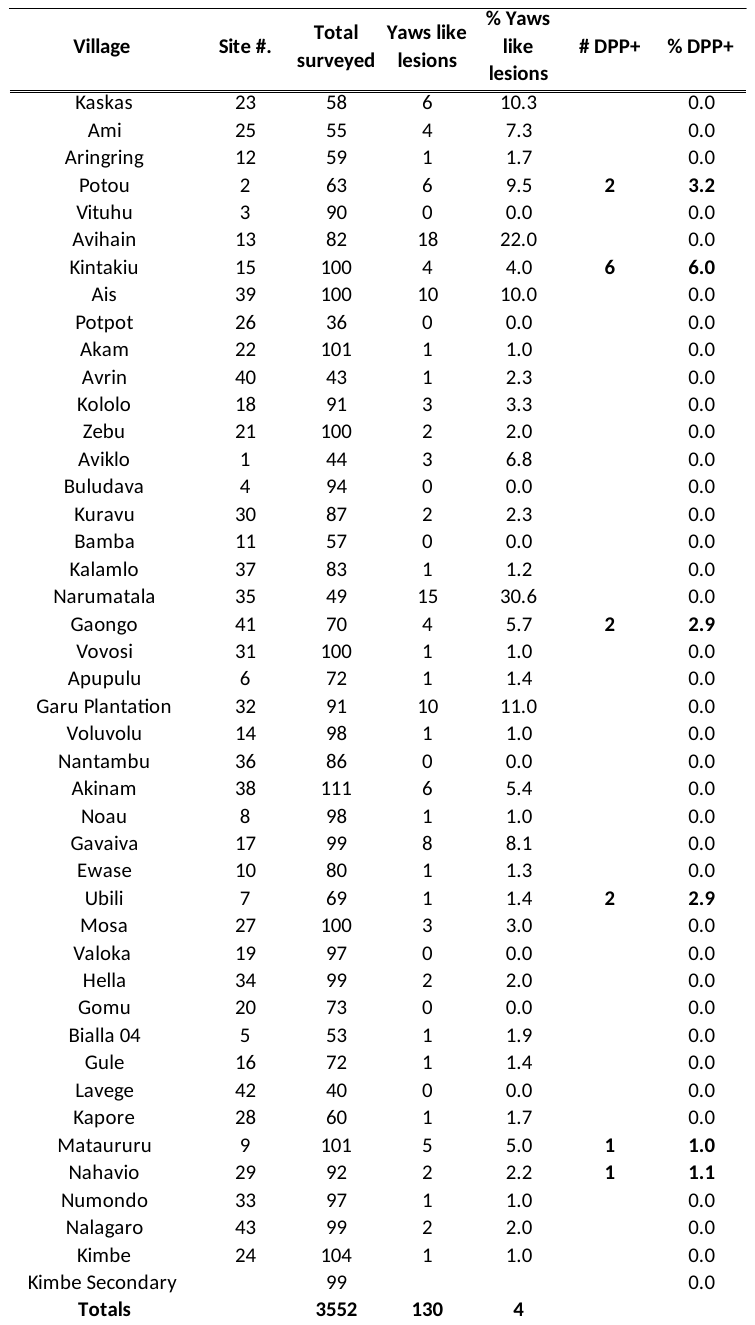

**S5 – Official coverage survey results after MDA 1, WNBP**

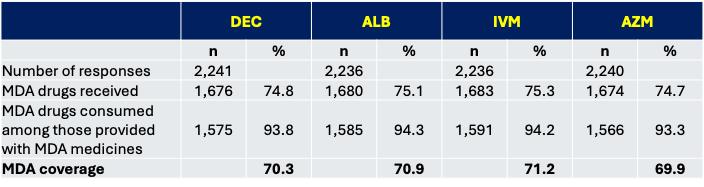
